# Efficacy and safety of erythropoietin for traumatic brain injury: a meta-analysis of randomized controlled trials

**DOI:** 10.1101/19006601

**Authors:** Motao Liu, Amy J. Wang, Gexin Zhao, Hua He, Ziv M. Williams, Kejia Hu

## Abstract

**Objective:** Recent studies regarding the effects of erythropoietin (EPO) for treating traumatic brain injury (TBI) have been inconsistent. This study conducts a meta-analysis of randomized controlled trials (RCTs) to assess the safety and efficacy of EPO for TBI patients at various follow-up time points.

**Methods:** A literature search was performed using PubMed, Web of Science, MEDLINE, Embase, Google Scholar and the Cochrane Library for RCTs studying EPO in TBI patients published through March 2019. Non-English manuscripts and non-human studies were excluded. The assessed outcomes include mortality, neurological recovery and associated adverse effects. Dichotomous variables are presented as risk ratios (RR) with a 95% confidence interval (CI).

**Results:** A total of seven RCTs involving 1197 TBI patients were included in this study. Compared to the placebo arm, treatment with EPO did not improve acute hospital mortality or short-term mortality. However, there was a significant improvement in mid-term (6 months) follow-up survival rates. EPO administration was not associated with neurological function improvement. Regarding adverse effects, EPO treatment did not increase the incidence of thromboembolic events or other associated adverse events.

**Conclusions:** This meta-analysis indicates a slight mortality benefit for TBI patients treated with EPO at mid-term follow-up. EPO does not improve in-hospital mortality, nor does it increase adverse events including thrombotic, cardiovascular and other associated complications. Our analysis did not demonstrate a significant beneficial effect of EPO intervention on the recovery of neurological function. Future RCTs are required to further characterize the use of EPO in TBI.

## Introduction

Traumatic brain injury (TBI) is a major cause of death and lifelong disability around the world, and predominantly affects younger and middle-aged people^1^. The United States Center for Disease Control estimates that TBI results in more than 280,000 hospitalizations and 2.2 million emergency department visits, and contributes to the deaths of over 52,000 people annually^2^. As the direct and indirect health-care costs of TBI are estimated at more than $76.5 billion^3^, TBI is a pressing medical and public health problem.

The mechanisms of TBI are typically divided into “primary” and “secondary” injury. Primary injury refers to the direct trauma to the brain, while secondary injury^4^ refers to the sequelae, which consists of a complex set of cellular and molecular processes inducing destruction of mitochondrial integrity, progressive neuronal loss through necrosis and apoptosis, accumulation of lactate, and cytotoxic swelling of cells. They will reduce cerebral perfusion by causing brain edema and an increase in intracranial pressure. These deteriorations can occur for days, weeks and even months following the initial trauma, resulting in delayed tissue damage^5^.

Over the past few decades, our understanding of the dynamic TBI pathophysiology has improved significantly^6^. More than 100 compounds are currently being investigated in preclinical studies for the treatment of secondary injury. However, almost all Phase II/III TBI clinical trials have failed^7^.

Erythropoietin (EPO), a hemopoietin growth factor with neurocytoprotective effects from the type 1 cytokine superfamily, has been proved to be a promising neuroprotective therapeutic agent in a variety of neurological injuries including TBI^8^. In experimental models, EPO has been proved to stimulates hematopoiesis, and possess neuroprotective and neuroregenerative effects through reduction of apoptosis, relieve inflammation, oxidative stress, and excitotoxicity^9^. A meta-analysis about the effect of EPO in experimental TBI in animal models concluded that EPO could reduce the lesion volume and improve neurobehavioral outcomes, which might be beneficial in treating experimental animal modes of TBI^10^. However, EPO’s mechanism of action was only partially understood through laboratory experiments, and the potential benefits and possible risks of EPO for TBI patients still need investigation. Pharmaceutical therapy with net clinical benefit had been lacking^11^. Clinical evidence of the EPO therapy quickly evolved but conveyed conflicting results^12^.

One prior systematic review on EPO for TBI by Liu et al. (2016)^13^ suggests that EPO reduces overall mortality and shortens hospitalization time without increasing the risk of DVT, but does not improve favorable neurological outcomes. While this study represents an important first step in interpreting the EPO for TBI literature, its investigation would have been strengthened by the inclusion of additional pertinent RCTs, analyses of multiple follow-up periods, and analysis of additional outcome measures.

Therefore, the current meta-analysis of RCTs aims to analyze a broader set of outcomes and adverse events and to validate the efficacy, functional outcomes, and safety of EPO treatment for TBI patients.

## Materials and Methods

We followed the guidelines proposed by the Cochrane Collaboration in the *Cochrane Handbook for Systematic Reviews of Interventions* (http://www.cochrane-handbook.org) and the recommended Preferred Reporting Items for Systematic Reviews and Meta-Analyses (PRISMA) statement to report this meta-analysis. The Institutional Review Board (IRB) approval was not required^14^.

### Search strategy

We conducted a systematic literature search (on March 1, 2019) of PubMed, EMBASE and the Cochrane Central Register of Controlled Trials for RCTs evaluating the efficacy or safety of EPO for TBI. We also searched the ClinicalTrials.gov registry, supplemented by manual searches of bibliographies of key retrieved articles and relevant reviews for additional published and unpublished data. We further checked the search engine “Google” for abstracts, conference proceedings, or unpublished studies. We used a combination of keywords and exploded medical subject headings (MeSH) including “EPO”, “erythropoietin” and “TBI”, “traumatic brain injur*”, “brain injur*”, “head injur*”. Another search using the same strategy was conducted on April 10, 2019, to identify additional publications.

### Inclusion criteria

We included RCTs if they met the following criteria: (1) Population: patients with a diagnosis of TBI; (2) Intervention: received intravenous or subcutaneous injection of EPO; (3) Comparison: placebo with no treatment; (4) Outcome measures: the primary outcome was mortality rate (while inpatient, and at short- and mid-term follow-ups defined as 10 weeks to 3 months follow-up and 6 months follow-up respectively), the second outcome was favorable neurological outcome (defined as the proportion of patients who achieved a GOS score of 4 or GOS-E score of 5). Regarding treatment safety, we examined postoperative complications including incidence of any thromboembolic events (including deep venous thrombosis, pulmonary embolism, cardiac arrest, myocardial infarction), and other associated adverse events (including pneumonia, sepsis, seizure, gastrointestinal complications and incidence of RBC transfusion).

### Study selection

Two authors (intials and initials) independently removed the duplicate records and screened all titles and abstracts to identify potentially eligible studies. The full text of these potentially eligible studies were then obtained to further confirm the eligibility of the study for the meta-analysis. Any disagreements regarding eligibility were arbitrated by consensus with the help of a third reviewer (initials).

### Data extraction

The two authors independently extracted data from included trials, using a standard abstraction form of excel sheet. From each study we extracted the following items: the first author, year of publication, country where the study was done, study design, number of participants, sex, age, number of cases, clinical settings, intervention, time from intervention to treatment, and comparison arm. When the raw data were not available in the publications, we searched the supplemental attachments or contacted the authors of the original reports. All data extraction in duplicate numbers were excluded and discrepancies between the two reviewers were discussed until agreement was reached.

### Quality assessment

Two reviewers independently assessed the risk of bias in each of the included studies without being blinded to the authors, institutions, or manuscript journals. The eligible studies were evaluated using the Cochrane Collaboration’s Tool and according to the predefined checklist of the Cochrane Database of Systematic Reviews^15^. This checklist assesses the risk of bias in random sequence generation, allocation concealment, blinding of participants and personnel, blinding of outcome assessment, completeness of outcome data, degree of selective reporting, as well as other biases. Any discrepancies between the two reviewers were solved by consensus or involvement of the other reviewers.

### Statistical analysis

For dichotomous and continuous outcomes, the differences between the experimental and control arms were quantified as risk ratios (RR) and mean differences (MD) with *P* values and 95% confidence intervals (CIs). To assess the presence of heterogeneity, we used a formal Cochran Q test and quantified with the *I*^2^ statistic^16^. We considered heterogeneity to be mild if the *I*^2^ value was ≤ 50%. We used fixed-effects meta-analyses to combine results when *I*^2^ ≤ 50%. Otherwise, we applied a random-effects model. Potential for publication bias was assessed with the funnel plot and Egger regression test^17^. All statistical analyses were performed using Review Manager Software (RevMan version 5.3; Nordic Cochrane Centre, Cochrane Collaboration) and Stata Statistical Software (Stata 14.0, Stata Corp, College Station, Texas).

## Results

### Study selection and characteristics

The PRISMA statement flowchart (Figure 1) shows the detailed process of literature screening, study selection, and reasons for exclusion. Ultimately, 1197 TBI patients (611 treated with EPO, 586 treated with placebo) from seven RCTs were included in this meta-analysis.

**Figure 1.**
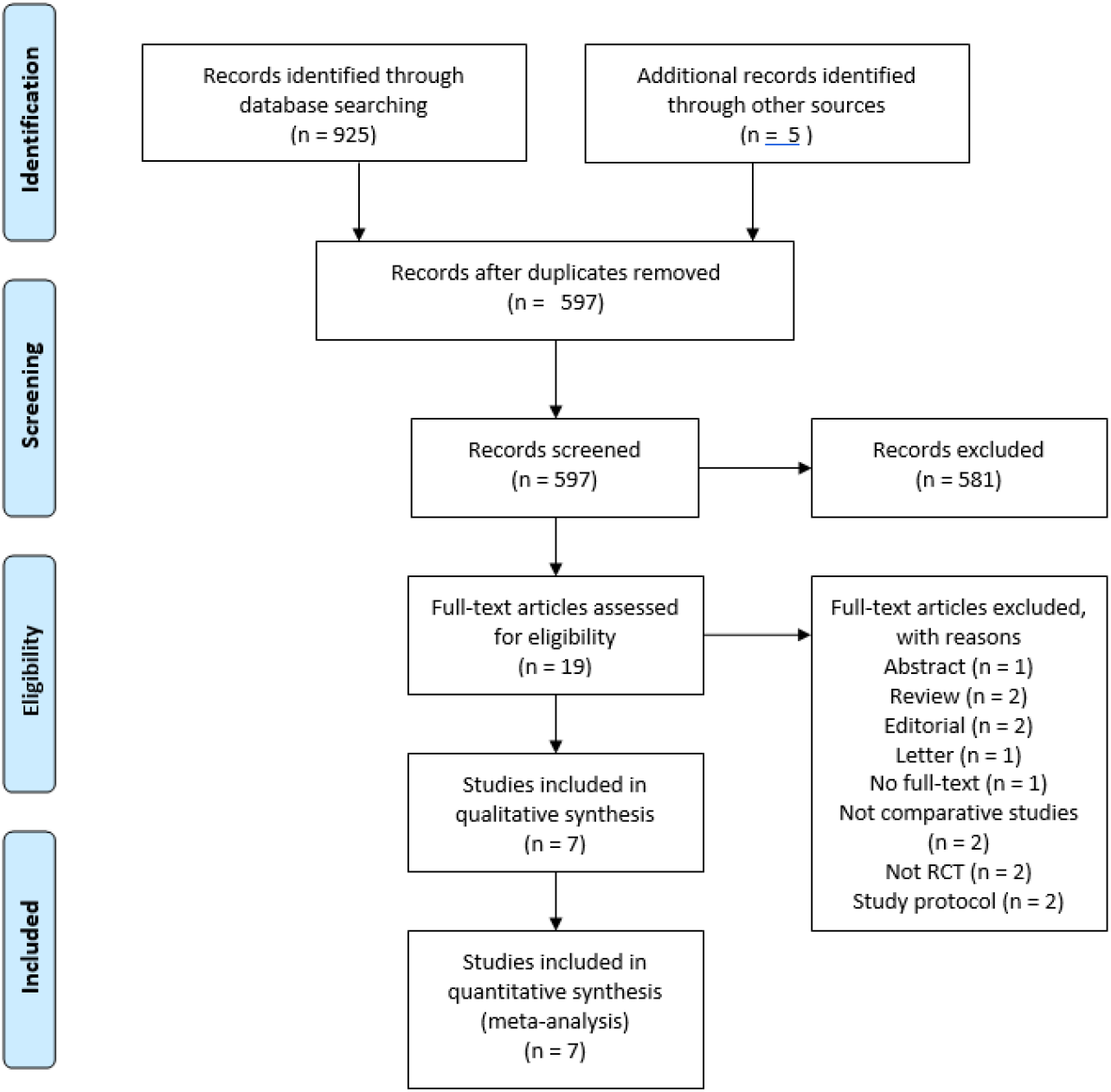
PRISMA flow diagram summarizing search strategy and selection of RCTs for the meta-analysis. RCT = randomized controlled trial.

Of these seven included RCTs, six studies utilized a normal saline placebo control and one study did not mention the use of placebos^18^. The mean age of participants ranged from 26.3 to 43.8 years. The time from trauma to intervention was within 6 to 24 hours; different EPO regimens and dosages were used: five studies injected EPO, rhEPO, or epoetin alfa subcutaneously while two gave EPO intravenously with the total dosage ranging from 12,000 international unit(IU) to 120,000 IU, and the EPO administration mostly started within six hours except one started in 24 hours^19^. In our study, Nichol *et al*. set a 24-h time window for recruitment, which was at odds with other included RCTs. The major characteristics of the seven RCTs are shown in Table 1, with slightly different enrollment criteria of each. ^20^

**Table 1:**
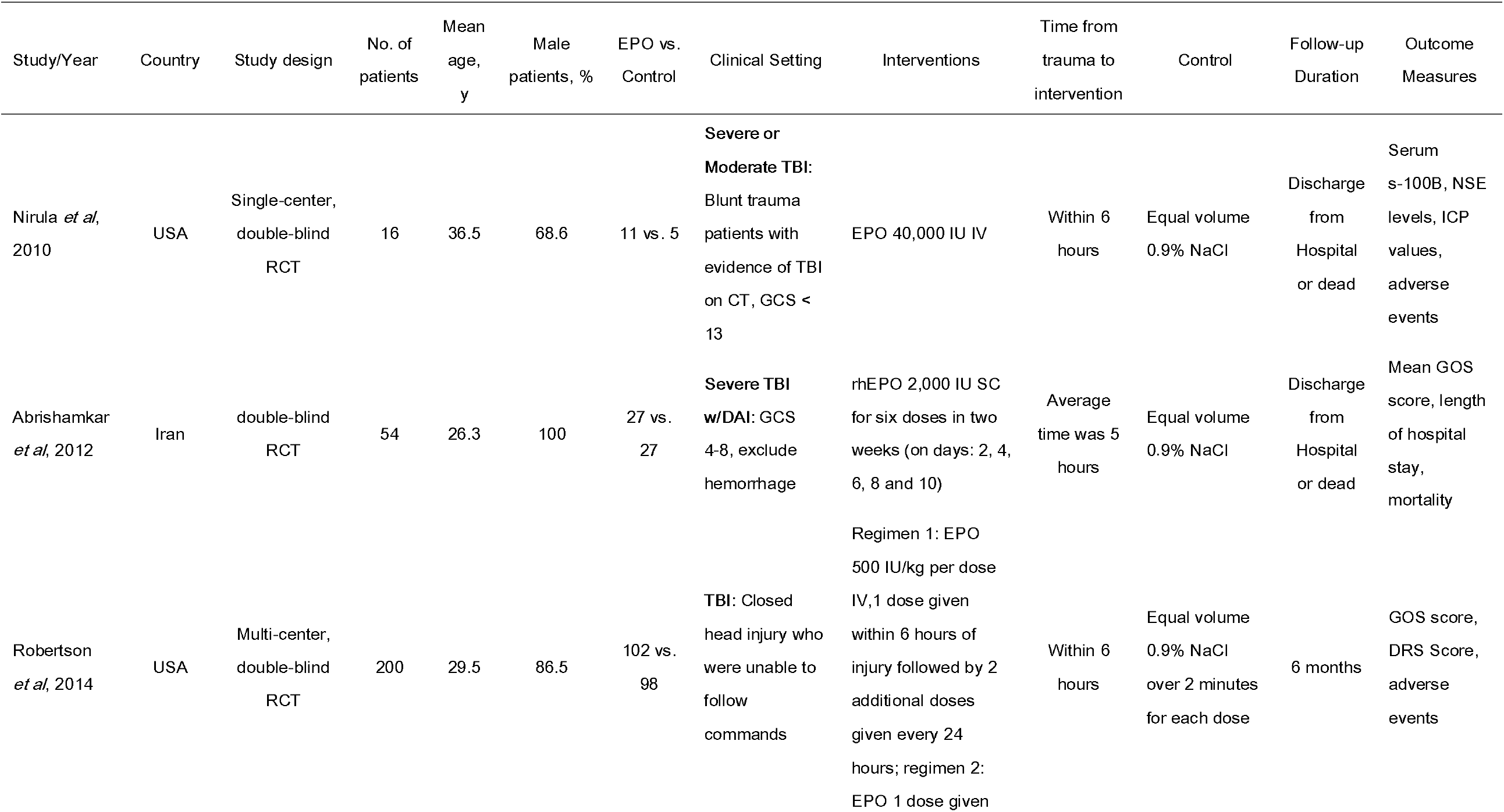
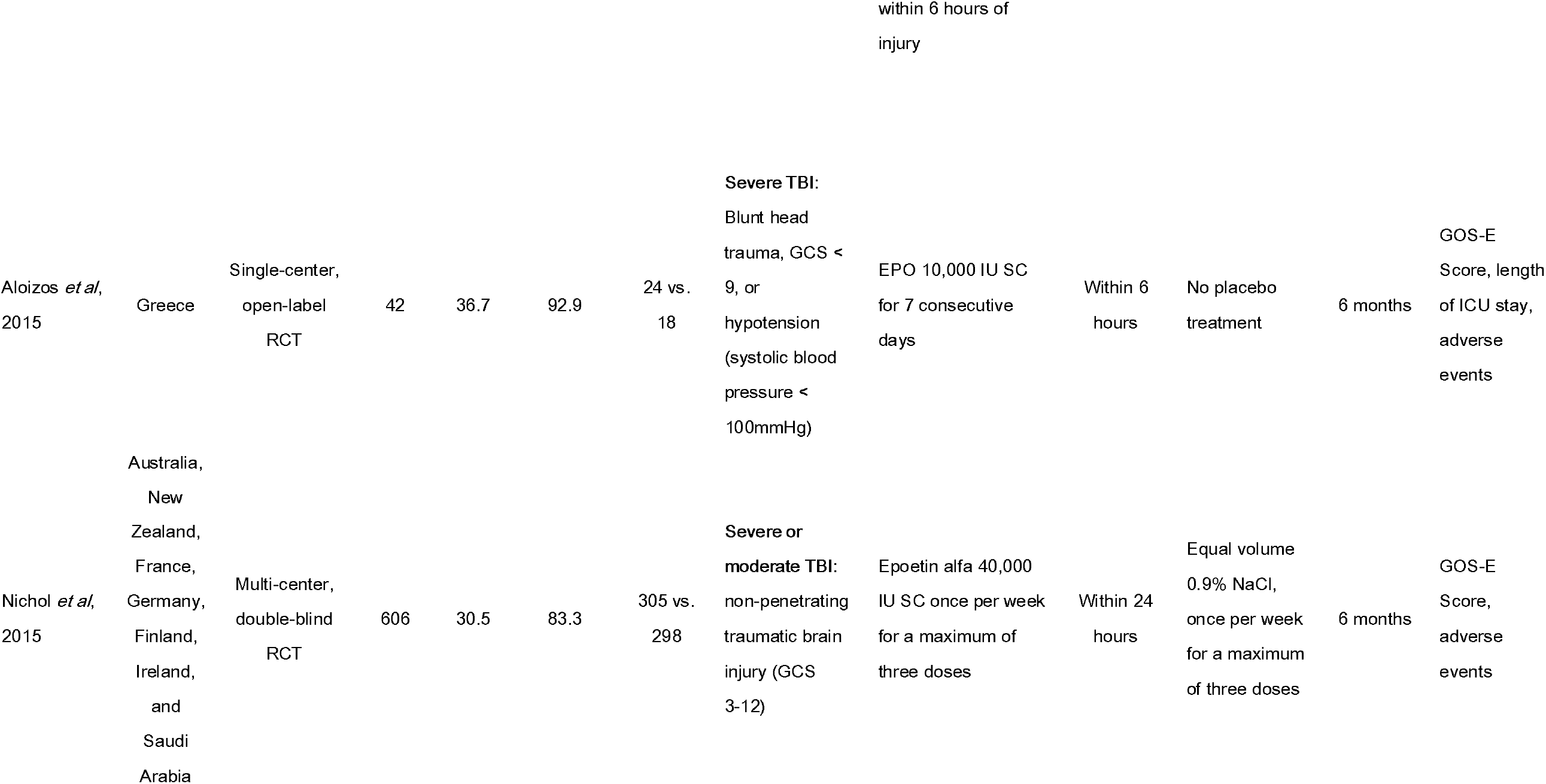
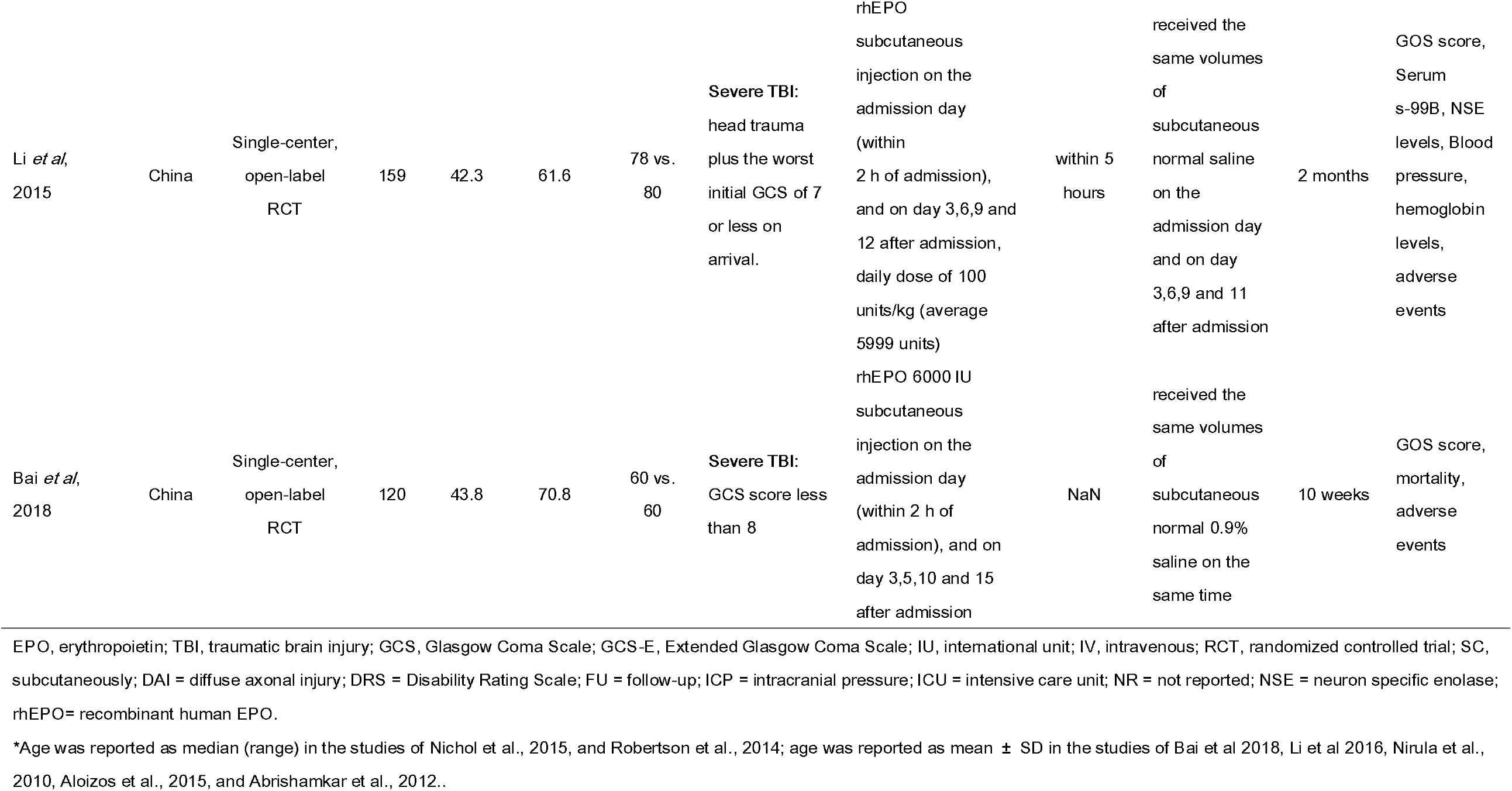
Characteristics of randomized controlled trials included in the meta-analysis

### Quality Assessment

Our assessments of each study’s risk of bias are summarized in Figure 2. One trial was determined to be at high risk for bias^18^, two trials were at low risk for bias^19 21^, and the other four were at unclear risk of bias^19 22-24^. Four studies describe their methods for generating the randomization sequence and for allocation concealment. Five studies reported blinding of participants and investigators, which might induce performance bias.

**Figure 2.**
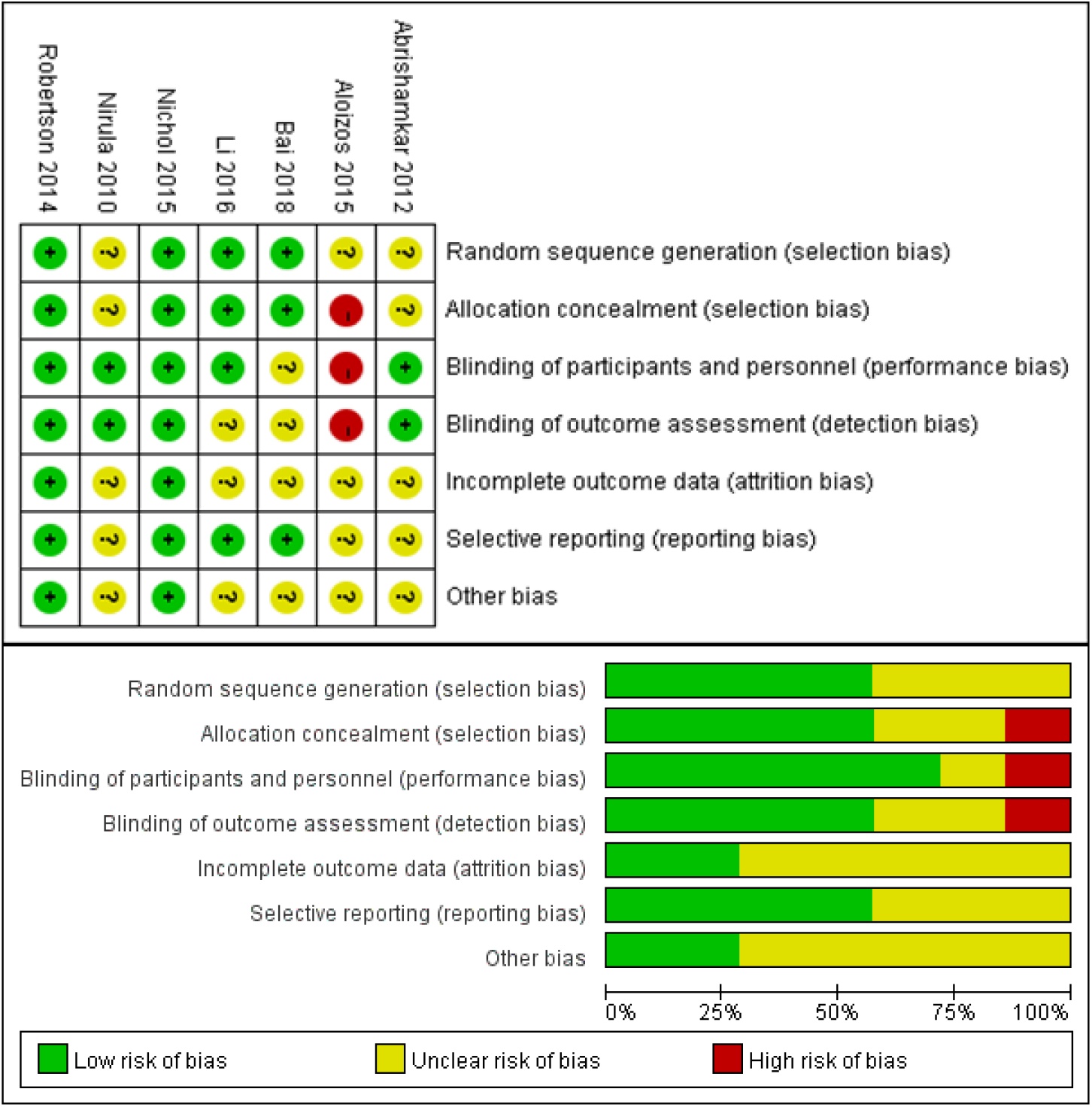
Risk of bias. Upper row: Each risk of bias metric for each included study. Lower row: Review authors’ judgements about each ‘Risk of bias’ item presented as percentages across all included studies. The overall risk of bias is relatively low. “+” indicates yes; “-” indicates no; “?” indicates not clear.

### Meta-analysis outcomes

#### Mortality

All studies reported on mortality (Figure 3). Compared to the placebo arm, there was a no significant reduction in acute hospital mortality (EPO:7.1%;placebo:9.2%,RR = 0.71, 95% CI [0.32–1.58]; *P* = 0.41) and short-term mortality (RR = 0.64, 95% CI [0.26–1.60]; *P* = 0.34) in the patients with TBI (RR = 0.69, 95% CI [0.50–0.95]; *p* = 0.03). Our analysis of mid-term mortality(six months follow-up) included a total of 792 patients (74.5% of all included patients, drawn from the Robertson et al. 2014 and Nichol et al. 2015 studies). Here, the EPO-treated group showed a significantly lower mortality (EPO:11.2%,placebo:16.4%,RR = 0.69, 95% CI [0.48–0.98]; *p* = 0.04), the mid-term result of EPO on mortality was in accordance the overall results.

**Figure 3.**
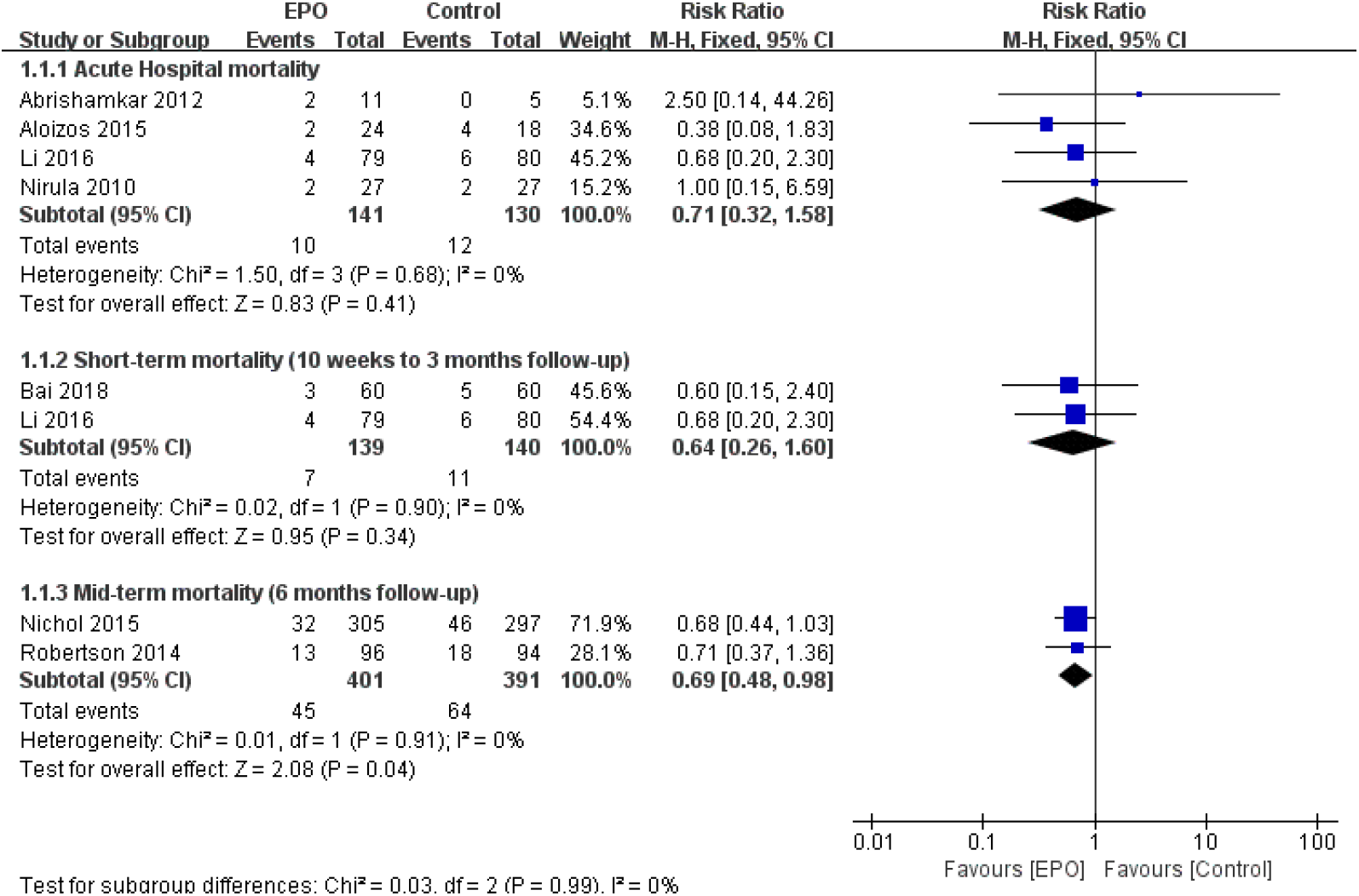
Effect of EPO intervention on mortality compared with control treatment at varying lengths of follow-up. Results are shown using a fixed-effect model with Risk ratio and 95% CIs. CI, confidence interval; M-H, Mantel-Haenszel.

#### Efficacy of EPO on Neurological Recovery

The neurological outcomes of TBI patients were evaluated by GOS or GOS-E varying from 10 weeks to 6 months (Figure 4). only Li et al had showed EPO significant better control group. However, EPO was not associated with favorable neurological function improvement GOS-E 5-8 (RR = 1.22, 95%CI [0.82–1.81]; *P* = 0.33).

**Figure 4.**
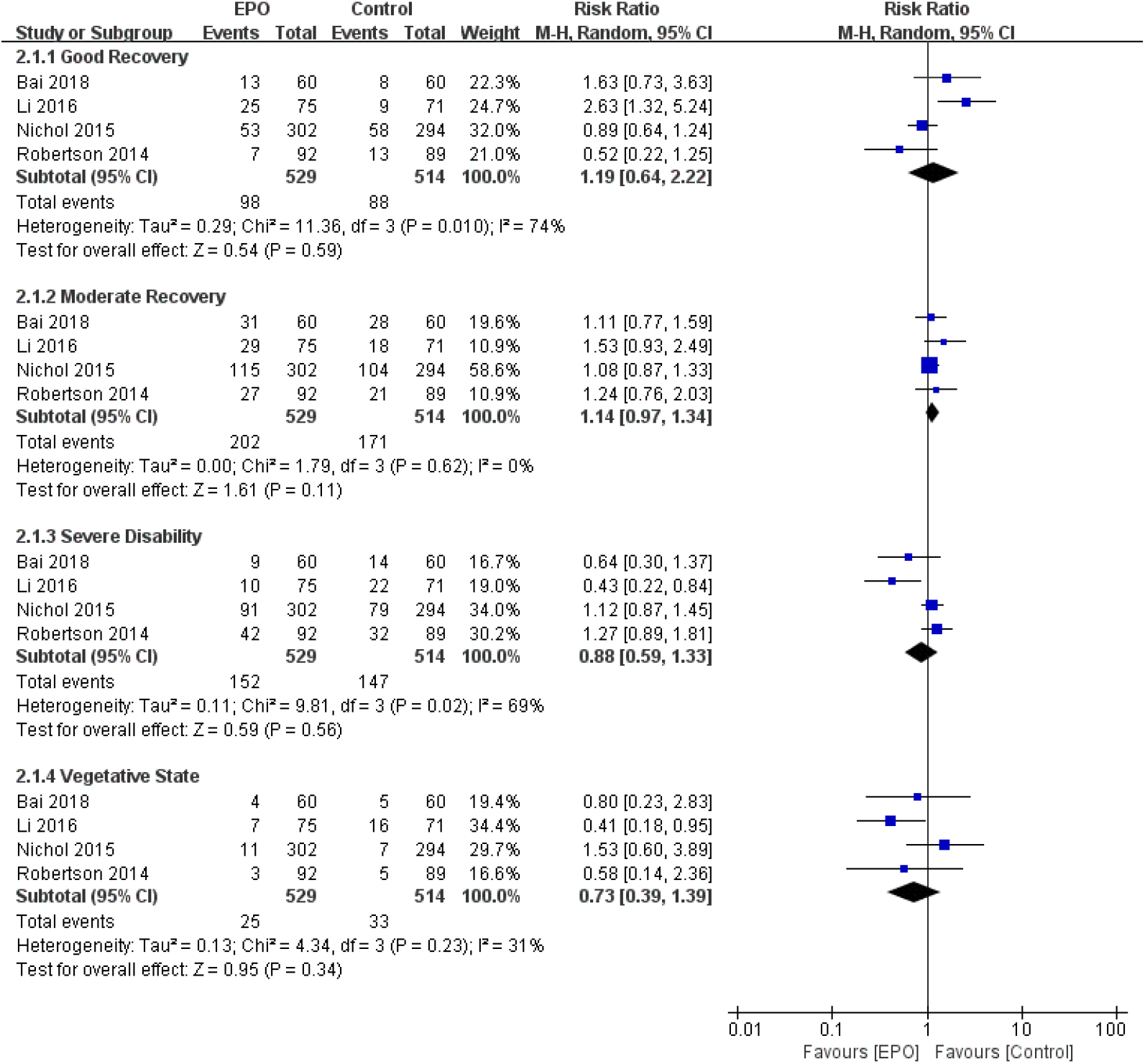
Effect of EPO intervention on neurological function recovery compared with control treatment for different levels according to GOS or GOS-E system at mid-term follow-up period. Results are shown by using a Random-effect model with Risk ratio and 95% CIs. CI, confidence interval; M-H, Mantel-Haenszel.

### Safety outcomes

#### Thrombotic and cardiovascular events

Five studies reported the incidence of thromboembolic events^15,17-19^ at the end of follow-up (N = 1980). Compared to the placebo treatment, EPO therapy did not increase the incidence of total thromboembolic events (EPO:19.9%,placebo:22.2%,RR = 1.01, 95% CI [0.65–1.57]; P = 0.97) in TBI patients (Figure 5). Our analysis of the pooled results demonstrate that there was no difference in rates of deep venous thrombosis (EPO:13.6,placebo:14.1%,RR = 0.98, 95% CI [0.45–2.14]; *P* = 0.96), pulmonary embolism (EPO:4.1%,placebo:3.3%,RR = 1.23, 95% CI [0.63–2.40]; *P* = 0.54), cardiac arrest (EPO:2.5%,placebo:1.8%,RR = 1.39, 95% CI [0.53–3.61]; *P* = 0.50), or myocardial infarction (EPO:0.9%,placebo:0.7%,RR = 1.22, 95% CI [0.29–5.07]; *P* = 0.79), (Figure 5).

**Figure 5.**
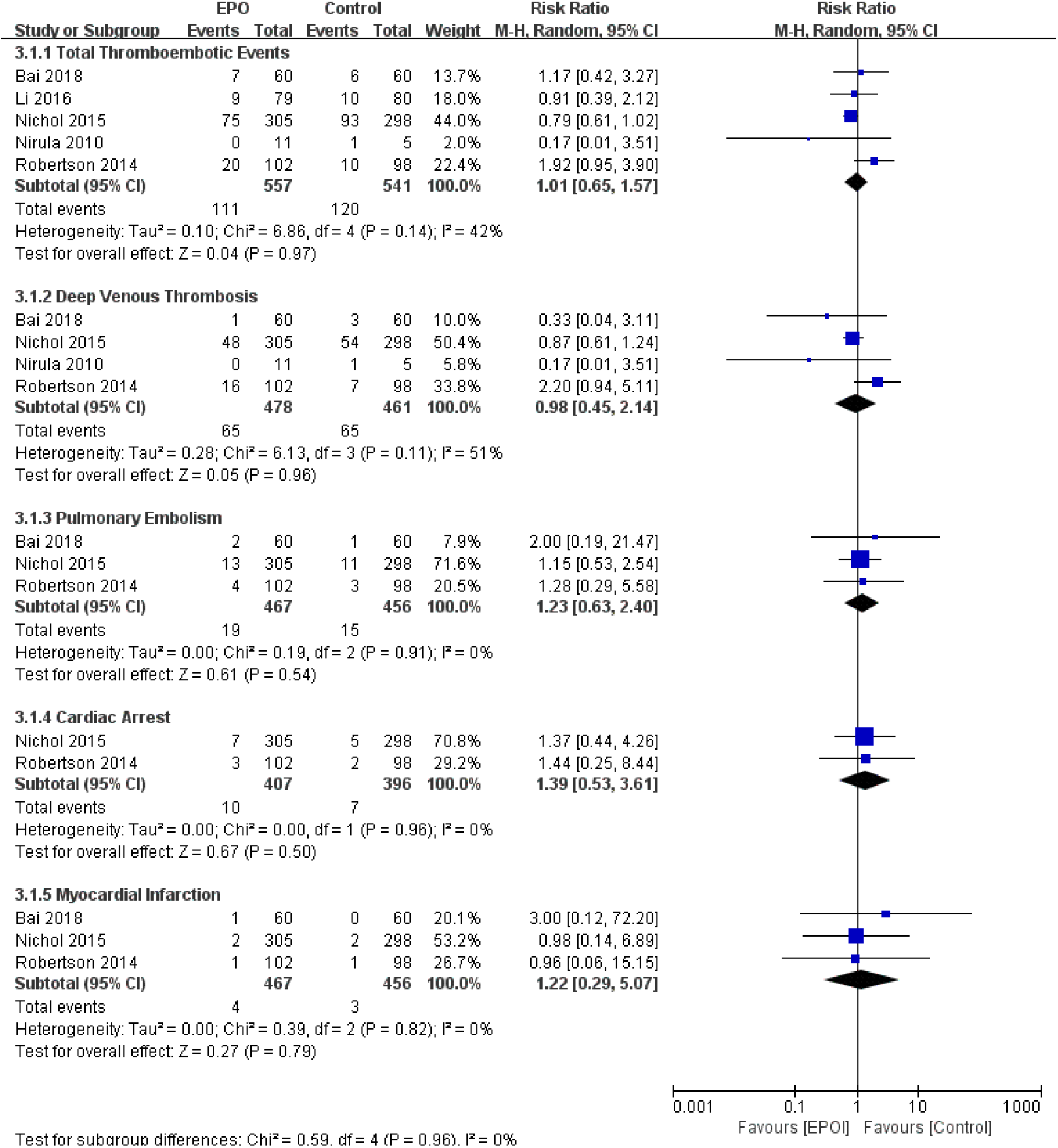
The thromboembolic adverse events of EPO intervention compared with control treatment at the end of follow-up period. Results are shown by using a Random-effect model with Risk ratio and 95% CIs. CI, confidence interval; M-H, Mantel-Haenszel.

#### Other associated adverse events

Our pooled results showed no differences regarding other adverse events between the EPO and control groups (detail shown in Figure 6). There were no differences in rates of pneumonia (EPO:10.8%,placebo:8.7%,RR = 1.25, 95%CI [0.73–2.15]; P = 0.42), sepsis (EPO:4.6%,placebo:3.5%,RR = 1.30, 95%CI [0.72–2.32]; P = 0.38), RBC transfusion (EPO:43.2%,placebo:45.7%,RR = 0.94, 95%CI [0.81–1.10]; P = 0.46), seizure (EPO:4.5%,placebo:3.7%,RR = 1.20, 95%CI [0.64–2.25]; P = 0.44), gastrointestinal complications (EPO:8.4%,placebo:7.5%,RR = 1.10, 95%CI [0.74–1.64]; P = 0.63). (Figure 6).

**Figure 6.**
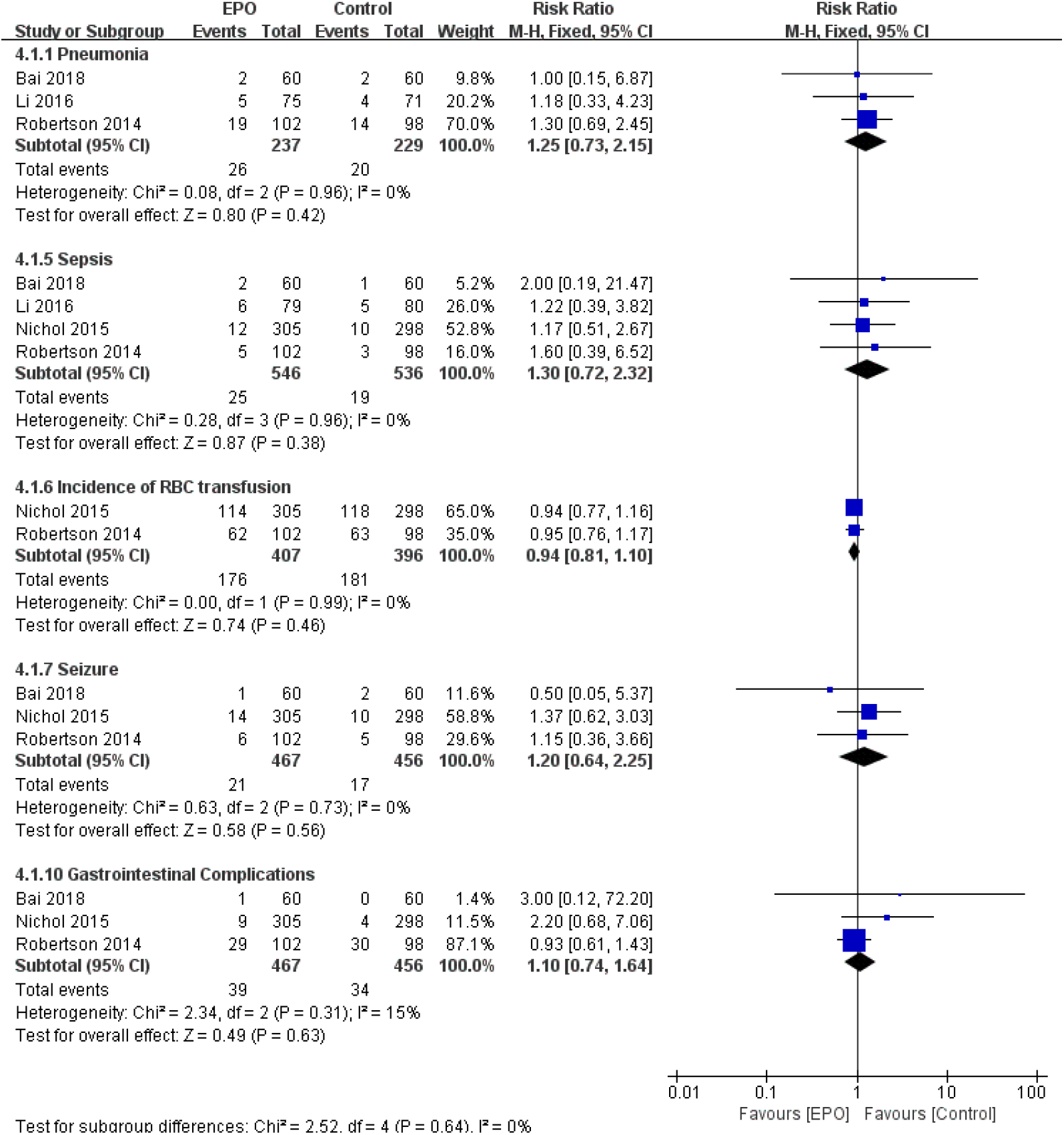
Other associated adverse events of EPO intervention compared with control treatment at the end of follow-up period. Results are shown by using a Random-effect model with Risk ratio and 95% CIs. CI, confidence interval; M-H, Mantel-Haenszel.

#### Publication bias

A funnel plot did not reveal any obvious asymmetry (Figure 7), and clear evidence of publication bias was not detected by Egger’s test.

**Figure 7.**
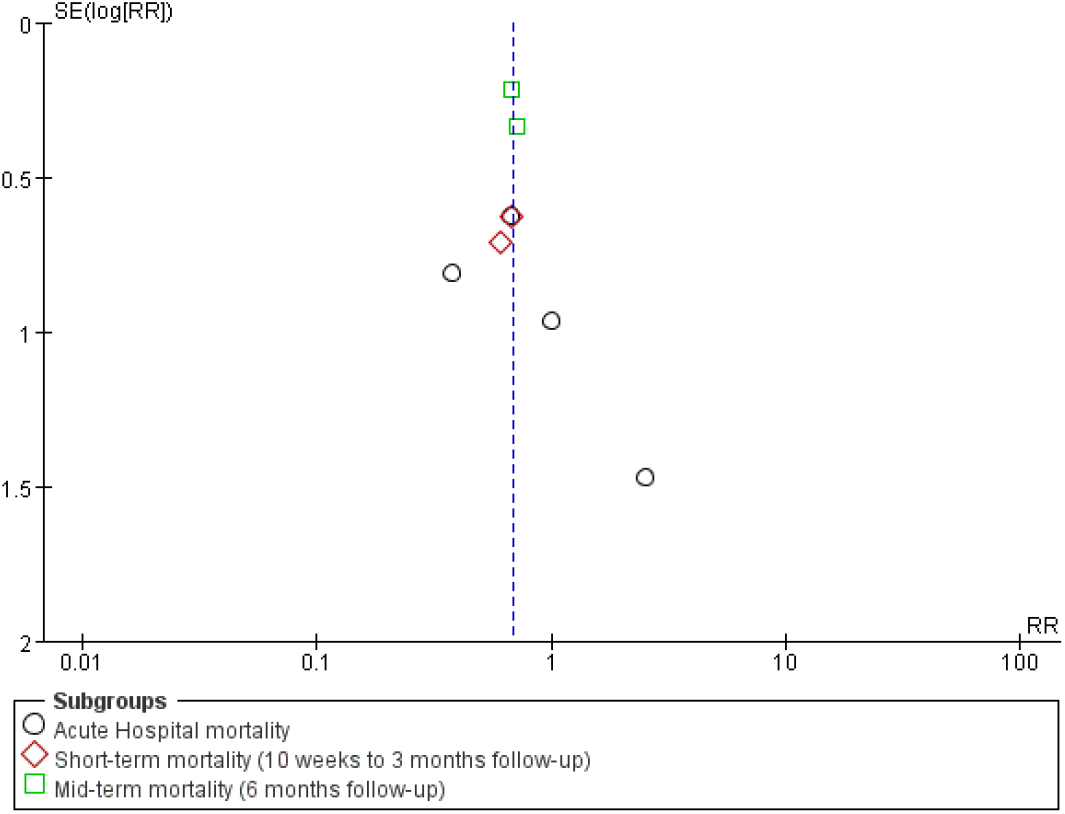
Funnel plot for the detection of publication bias. The funnel plot of pooled studies that evaluated the effects of EPO on mortality appears to be symmetrical.

## Discussion

### Principal findings

This meta-analysis demonstrates a survival benefit at 6 months follow-up to the administration of EPO to TBI patients. However, EPO intervention did not impact acute hospital mortality (in 10 weeks) or short-term mortality (10 weeks to 3 months). EPO therapy does not increase adverse events including thrombotic and cardiovascular complications. At the same time, EPO intervention had no significantly effect on the recovery of neurological function.

### Mortality

This current study of EPO treatment in TBI patients provides a novel analysis of survival rates at different follow-up times. The study was well-powered enough to detect a slight but significant six-month survival benefit with EPO treatment. While the concrete mechanism of mortality reduction in patients with TBI remains unclear, studies suggest that short-term mortality is related to either brain death or treatment withdrawal due to a perceived poor prognosis, whereas long-term mortality is believed to be mainly due to infection and multi-organ failure.^13^ Our results may suggest the TBI patients may benefit from EPO treatment for its long-term effect, which may relate to its organ-protective effects^25^.

### Neurological recovery

Preclinical laboratory studies suggest that EPO may decrease local tissue hypoxia in the brain, improve function of the blood-brain barrier, decrease cerebral edema, and attenuate secondary brain injury, making EPO theoretically well suited to treat TBI.^26^ However, our meta-analysis demonstrated no significant neurologic improvement in TBI patients following EPO treatment. This is concordant with Liu et al’s conclusion ^13^. Furthermore, our study further demonstrated that no difference at each subgroup level of GOS and GOS-E systems(RR = 1.22, 95%CI [0.82–1.81]; *P* = 0.33)..

The value of laboratory experiments for predicting the effectiveness of treatment strategies in clinical trials remains controversial. Several factors may contribute to the unguaranteed benefits in human trials compared with EPO being used in animal models. First, the characteristics of the experimental TBI tend to be simple and replicate only single factors in laboratory models^27^. In contrast, TBI in humans can result from a variety of etiologies such as the neglect of motorists, cyclists, construction workers and industrial workers in observing safety precautions, resulting in heterogeneous damage patterns including cranial fractures, intracerebral hemorrhage, cerebral contusion, cerebral edema, and soft-tissue injuries^28^. Second, there are differences between rodent and human systemic physiological and behavioral responses to neurotrauma, which may lead to different rates of survival as well as differences in neurological recovery^29^.

A broad spectrum of secondary events, complex cascade of molecular and cellular events is triggered by the initial injury. This all contribute to cell death and/or degeneration and worsen patient neurological outcomes but could, at least theoretically, be counteracted.^52^ To preserve and restore the integrity, function, and connectivity of the brain cells and improve the patient’s outcome, neuroprotective drugs should be administered as soon as possible and as long as the pathological cascades occur.^30^ An experimental study in mice suggested the importance of the therapeutic time-window within 6 hours after the initial TBI^31^. In laboratory studies, EPO can be administered as early as 5 minutes after injury^32^. While this short time to intervention is not always possible in the clinical setting, these studies suggest that TBI protocols should incorporate early recognition and diagnosis, and timely intervention. However, the dose and timing of EPO injections varied greatly across RCTs, and the current evidence is not strong enough to draw the conclusion that early intervention delivers better prognosis.^33^

### Complication

The EPO dose and therapeutic duration were not to reach a consensus, maybe due to the safety concerns of EPO has not been well established. Most of the evidence regarding the safety of EPO comes from its non-neurologic use; previous studies reported increased thromboembolic complications and/or mortality risks with EPO administration to cancer patients, critically-ill patients and patients with kidney disease^34^. One prior study administered EPO to acute ischemic stroke patients, which showed that EPO therapy significantly improved long-term neurological outcomes in patients after ischemic stroke, but the long-term recurrent stroke and mortality rate did not differ between the EPO-treated and placebo-control group.

Our finding suggests that use of EPO in TBI patients is safe and well-tolerated. However, the interactions between EPO and various physiologic variables as well as drugs commonly used in TBI patients are unknown. Future studies should further investigate the safety profile of EPO for TBI, especially when other commonly-used drugs involving in.

### Limitations and weakness

There are a number of limitations often inherent to meta-analyses that we encountered. First, the EPO treatment regimes differed across studies. The heterogeneity of the original RCTs may have reduced our ability to discern the true differences between the intervention and control arms. Second, the six-month mid-term follow-up time point is still a relatively brief time-window. Our analysis was limited by the varying follow-up durations of the included RCTs; further studies should conduct longer-term follow-up to continue coursing the efficacy and safety of EPO treatment for TBI. Finally, there are limited published data evaluating EPO treatment for TBI, publication bias was strongly suspected even though not detected.

### Future aim

Further exploration of molecular biomarkers should be anticipated to indicate the appropriate patients for EPO therapy after TBI^35^. Foundation of new appraisal system to assess the clinical effect is in demand, as with the originators of the GOS and GOS-E, survival is “an imperfect yardstick” in TBI. The current study did not demonstrate differences in neurologic recovery using the GOS and GOSE, but these scales are quite coarse and future studies should further investigate EPO as a neuroprotective intervention in TBI using more sensitive indicator suggested to detect the realistic slight, but still clinically meaningful, functional improvement. The time– and dose–response relationships of EPO treatment in TBI patients also needs to be better delineated. These aims can be accomplished with better homogenization of included patients, investigation of multiple dosages, standardization of intervention time, coupled with integrated multidimensional outcomes.

## Conclusions

This meta-analysis highlights the potential mid-term survival benefit of EPO treatment for TBI without increasing the risk of adverse events. However, further well-designed investigations of the effect of EPO in TBI patients are warranted to guide management.

## Data Availability

The data and software code that support the findings of this study are available from the corresponding author upon reasonable request.

## Abbreviations

CI: confidence interval
DVT: deep vein thrombosis
EPO: erythropoietin
GOS: Glasgow Outcome Scale
MD: mean difference
RCT: randomized controlled trial
RD: risk difference
RR: risk ratio
TBI: traumatic brain injury.

## Notes

### Competing Interest Statement

The authors have declared no competing interest.

### Funding Statement

None

### Author Declarations

All relevant ethical guidelines have been followed and any necessary IRB and/or ethics committee approvals have been obtained.

Any clinical trials involved have been registered with an ICMJE-approved registry such as ClinicalTrials.gov and the trial ID is included in the manuscript.

